# Modeling for Corona Virus Outbreak in IRAN

**DOI:** 10.1101/2020.03.24.20041095

**Authors:** Maryam Moghadami, Ka Wa, Mohammad Hassanzadeh, Aziz Hedayati

**Affiliations:** Knowledge Management, Faculty of Management and Economics, Tehran university, Tehran. Iran; Architecture & Environmental Engineering from the University of Nottingham, landan; Knowledge and Information science Dept, Faculty of Management and Economics Tarbiat Modares University Tehran. Iran; Head of Dept. Knowledge & InformationScience, Azarbaijan Shahid Madani University,Tabriz. Iran

## Abstract

**Background:** As the outbreak of coronavirus disease 2019 (COVID-19) is a worldwide pandemic, it is rapidly expanding in Iran, real-time analyses of epidemiological data are needed to increase situational awareness and inform interventions. In this study, we built a predictive model based on the cumulative trend of new cases and deaths for the top five provinces. we will also look at modeling the trends for confirmed cases, deaths and recovered for the whole country.

**Method:** In this study, we have chosen to apply the exponential smoothing model to iteratively forecast future values of a regular time seires from weightedaverages of past daily values of the series. This method is exponential because the value of each level is influenced by every preceeding actual value to an exponentially decreasing degree – more recent values are given a greater weight. The available data is too small to identify seasonal patterns and make predictable variation in value, such as annual fluctuation in temperature relative to the season. Trend is a tendency in the data to increase or decrease over time.

**Results:** If no control measures are put in place, it is expected that over 40,000 would be infected in Tehran around the middle of June. However, if control measures were implemented with a high degree of success, one would expect the spreadof the COV-19 virus would peak at the start of April with a downward trend dropping off by the end of May (70 days). In the scenario, that no further measures are implemented, one would expect the spread of COVID-19 to continue on a gentle incline, reaching 21,000 by mid-June. The same processhas been applied to review the confirmed, deaths and recovered dataset. The forecast has been carried out for the next 30 days, a shorter timeframe has been selected as there is a high probability that the Iranian New Year’s celebration, Farvardin, first month of Spring (30^th^ March in Western calendar) will have an impact on the infection rate following the event.

The best predictive model predicts the confirmed cases to be in the range of 35,000-70,000, with the number of reported COVDI-19 deaths to be between 3,000 – 5,000 and 5,000 – 30,000 of recovered cases.

**Conclusions:** Modeling outbreak of Covid-19 shows that the number of patients and deaths is still increasing. Contagious diseases follow an exponential model and the same be Haves this one. This is because, the virus can spread to others and finally each person turns into a carrier of the virus and transmit it to another person. Disease control depends on disconnection and social distancing. In addition, many factors are effective in stopping the disease. These include citizens’ participation in the prevention process, health education, the effectiveness of instructive traditions, environmental conditions, and so on.

---

On 31 December 2019, the World Health Organization(WHO) China Country Office was informed of cases of pneumonia of unknown etiology (unknown cause) detected in Wuhan City, Hubei Province of China, and WHO reportedthat a novel coronavirus (2019-nCoV),which was named as severe acute respiratory syndrome coronavirus 2 (SARS-CoV-2) by International Committee on Taxonomy of Viruses on 11 February, 2020, was identified as the causative virus by Chinese authoritieson 7 January(1)

During the <u>2019–20 coronavirus pandemic</u>, <u>Iran</u> reported its first confirmed cases of <u>SARS-CoV-2</u> infections on 19 February 2020 in <u>Qom</u>(2). As of 17 March 2020, according to Iranian health authorities, there had been 988 <u>COVID-19</u> deaths in Iran with more than 16,000 confirmed infections(3,4,5) This respiratory disease caused by a coronavirus is one of the leading causes for serious illnesses in people all over the world. According to the global statistics of fatalities caused by coronavirus, and its spread in Iran, it is vital and essential to forecast its outbreak by a model.

As the outbreak of coronavirus disease 2019 (COVID-19) is a worldwide pandemic, it is rapidly expanding in Iran, real-time analyses of epidemiological data are needed to increase situational awareness and inform interventions. Previously, real-time analyses have shed light on the transmissibility, severity, and natural history of an emerging pathogen in the first few weeks of an outbreak, such as with severe acute respiratory syndrome (SARS), the 2009 influenza pandemic, and Ebola. Analyses of detailed line lists of patients are particularly useful to infer key epidemiological parameters, such as the incubation and infectious periods, and delays between infection and detection, isolation, and reporting of cases. However, official individual patient data rarely become publicly available, when the information is most needed. This is an analysis of the COVID-19 out-break in Iran. In this population-level observational study, I used Iranian Ministry of Health reports downloaded from GitHub, an online datasharing platform. This dataset is updated on a daily basis with a 24 hour delay. The dataset include time-stamped counts of the daily cases and deaths within each province in Iran.

In this study, we will build a predictive model based on the cumulative trend of new cases and deaths for the top five provinces. I will also look at modeling the trends for confirmed cases, deaths and recovered for the whole country.

## Method

In this study, I have chosen to apply the exponential smoothing model to iteratively forecast future values of a regular time seires from weighted averages of past daily values of the series. This method is exponential because the value of each level is influenced by every preceeding actual value to an exponentially decreasing degree – more recent values are given a greater weight.

Due to the lack of historical data, seasonality analysis has been removed from the modelling and the trends are analysed based on the daily timeframe. The available data is too small to identify seasonal patterns and make predictable variation in value, such as annual fluctuation in temperature relative to the season. Trend is a tendency in the data to increase or decrease over time.

The predictive model will be tested against four regression model evaluations for robustness. They are as follow:

- MAE – mean absolute error, this gives less weight to the outliers
- MAPE – similar to MSE, but normalized by true observation. The downside is when true observation is zero, this metric could be problematic.
- MSE – mean squared error, it is like a combination measurement of bias ad variance of the prediction. For eample, bias squared add variance.
- RMSE – root MSE, this takes the root of MSE to bring the unit back to actual value. It is the standard deviation of residuals (prediction errors). Residuals are a measure of of how far from the regression line data points.

Both MAE & RMSE can range from 0 to infinity. They are negatively-oriented scores: lower values are better.

The Akaike Information Criterion, AIC test will be applied to the predictive model. This purpose of this test is to see how well the model fits the dataset without over-fitting it. The AIC score rewards models that achieve a high goodness-of-fit score ad penalizes them if they become over complex. A low AIC score indicates a better fit.

### Statistical analysis

Since the outbreak began in early February 2020, the rate of infection and number death has increased significantly as seen in the illustration below. On the 19^th^ February, the first 2 deaths were reported, on the 23^rd^ February, 4 additional province reported COVID-19 deaths, and within the space 28 days the virus has spreaded across the whole country (see illustration 2), by 20^th^March 2020, Tehran, the capital city of Iran reported close to 4000 confirmed cases of COVID-19.

**Illiustration 1.**
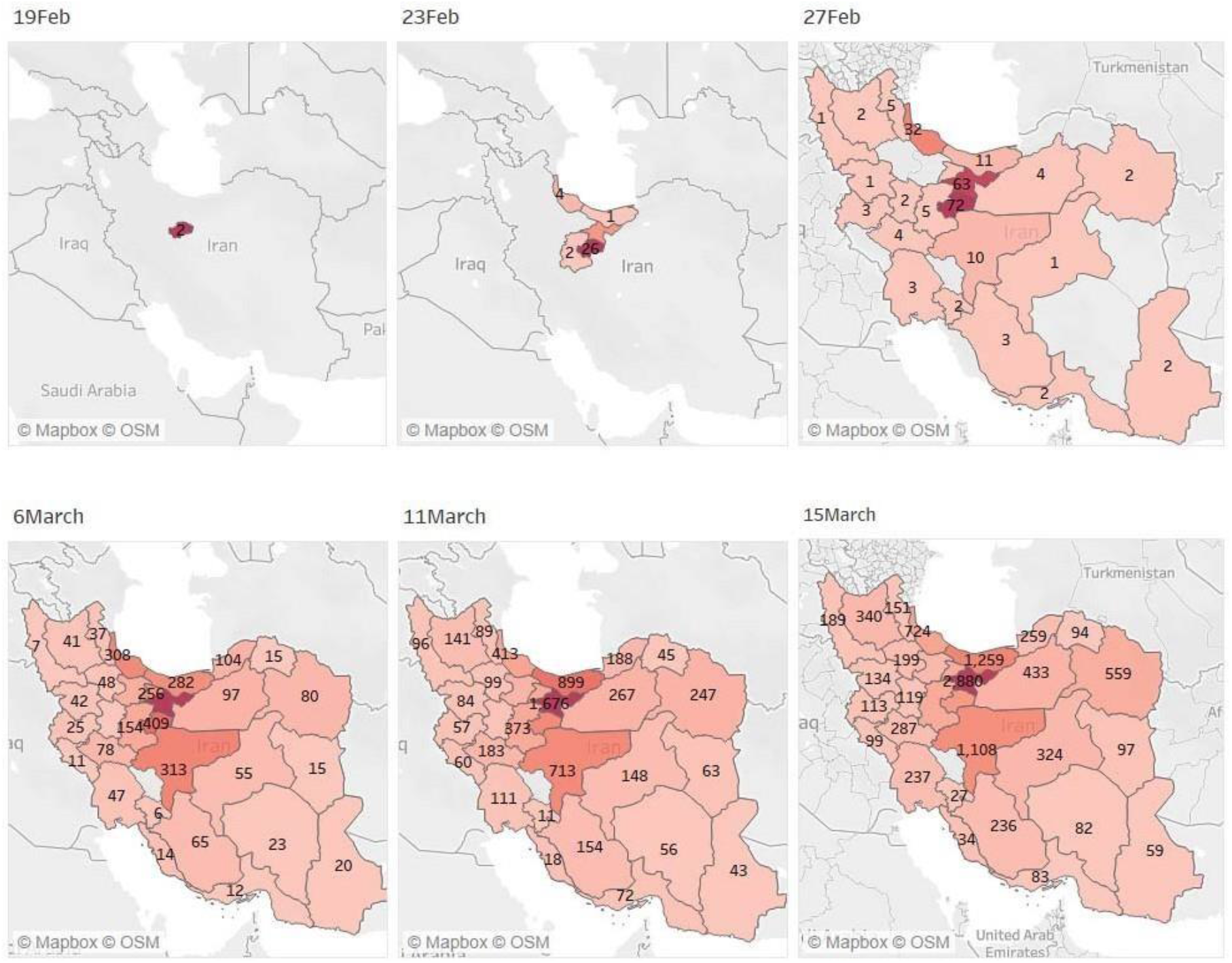

**Illustration 2.**
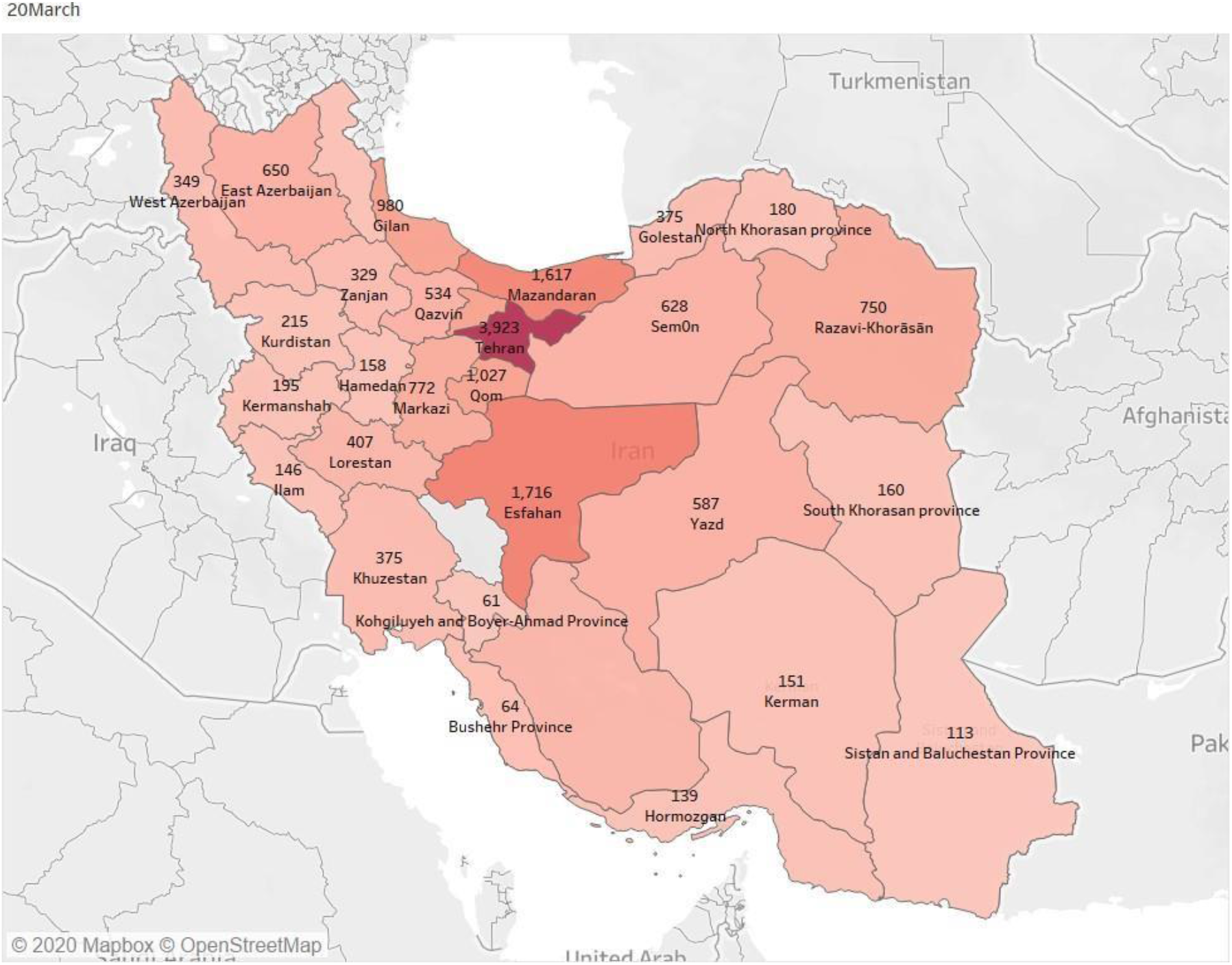

**Illustrattion 3.**
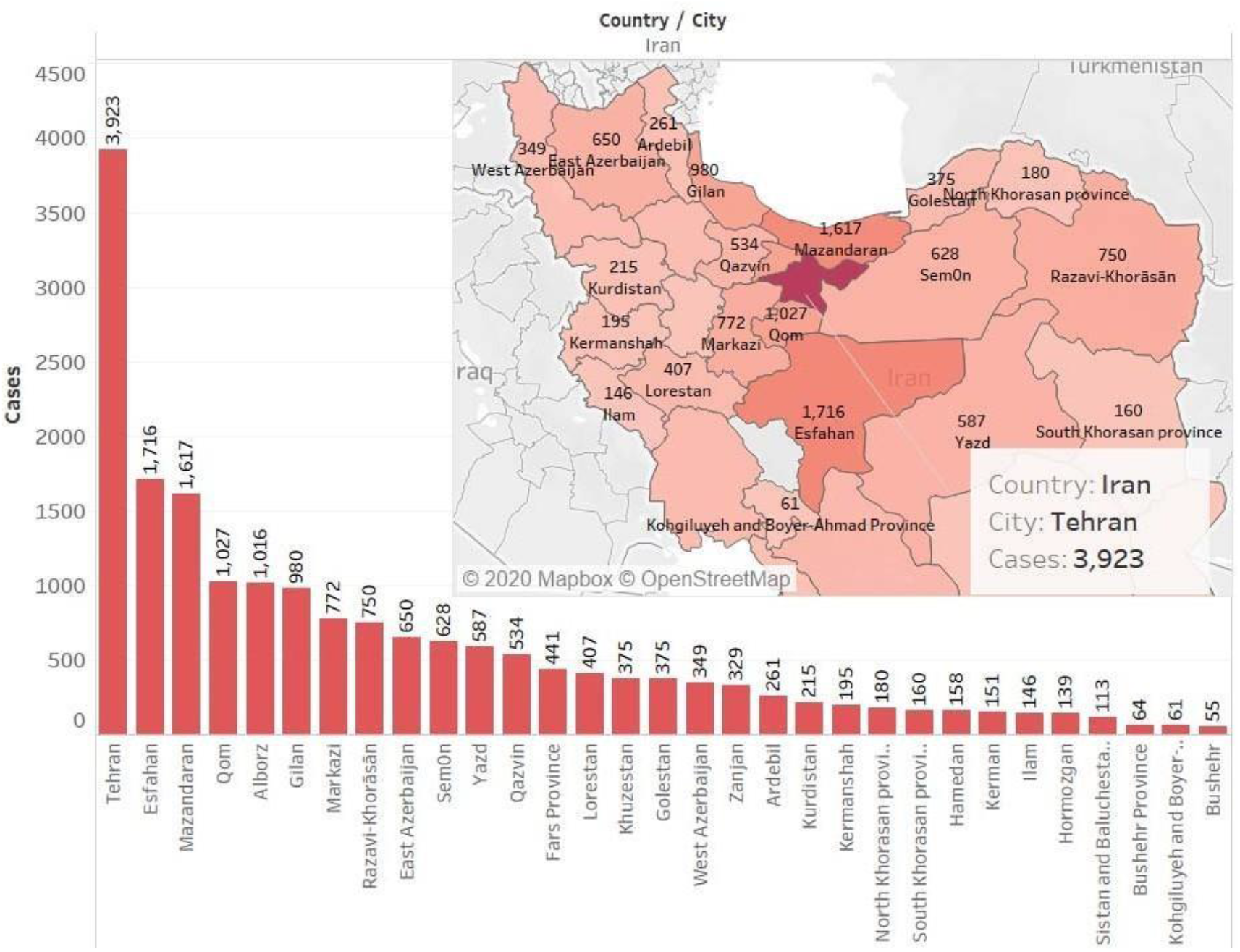

The total number of confirmed cases of infected patients on the 20^th^ March 2020 is 19,661, with 1,411 deaths and 6,226 recovered. The top 10 most affected provinces are Tehran, Esfahan, Mazandaran, Qom, Alborz, Gilan, Markazi, Razavi-Khorasan, East Azerbaijan and Semon, which can be reviewed in illustration 3, province ranked by confirmed cases from the highest number to the least.

**Illustration 4.**
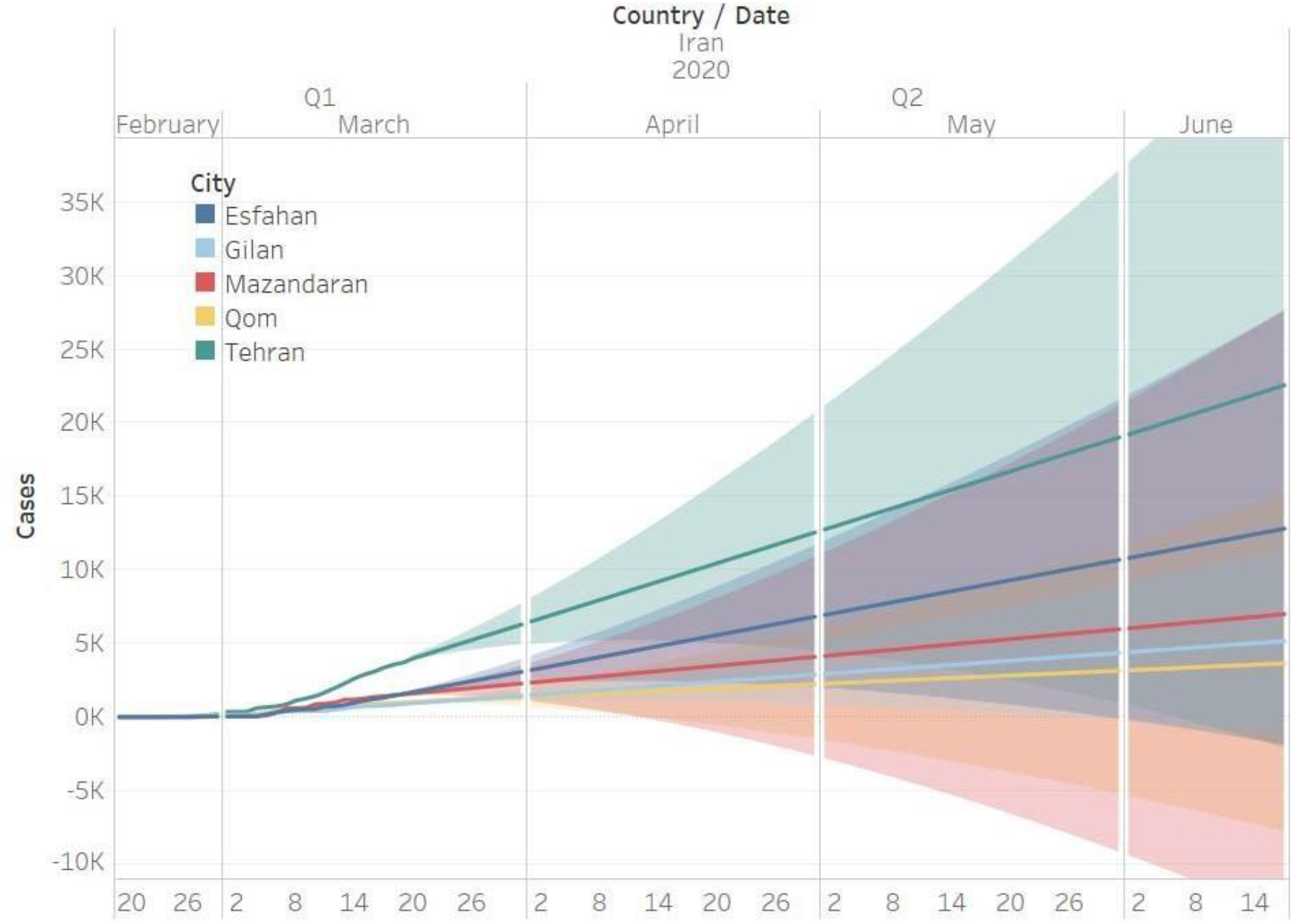

Based on the exponential smoothing model, the projection for the next 90 days across the top 5 with the most reported confirmed cases are shown in illustration 4. If no control measures are put in place, it is expected that over 40,000 would be infected in Tehran around the middle of June. However, if control measures were implemented with a high degree of success, one would expect the spread of the COV-19 virus would peak at the start of April with a downward trend dropping off by the end of May (70 days). In the scenario, that no further measures are implemented, one would expect the spread of COVID-19 to continue on a gentle incline, reaching 21,000 by mid-June.

**%Iluustration 5.**
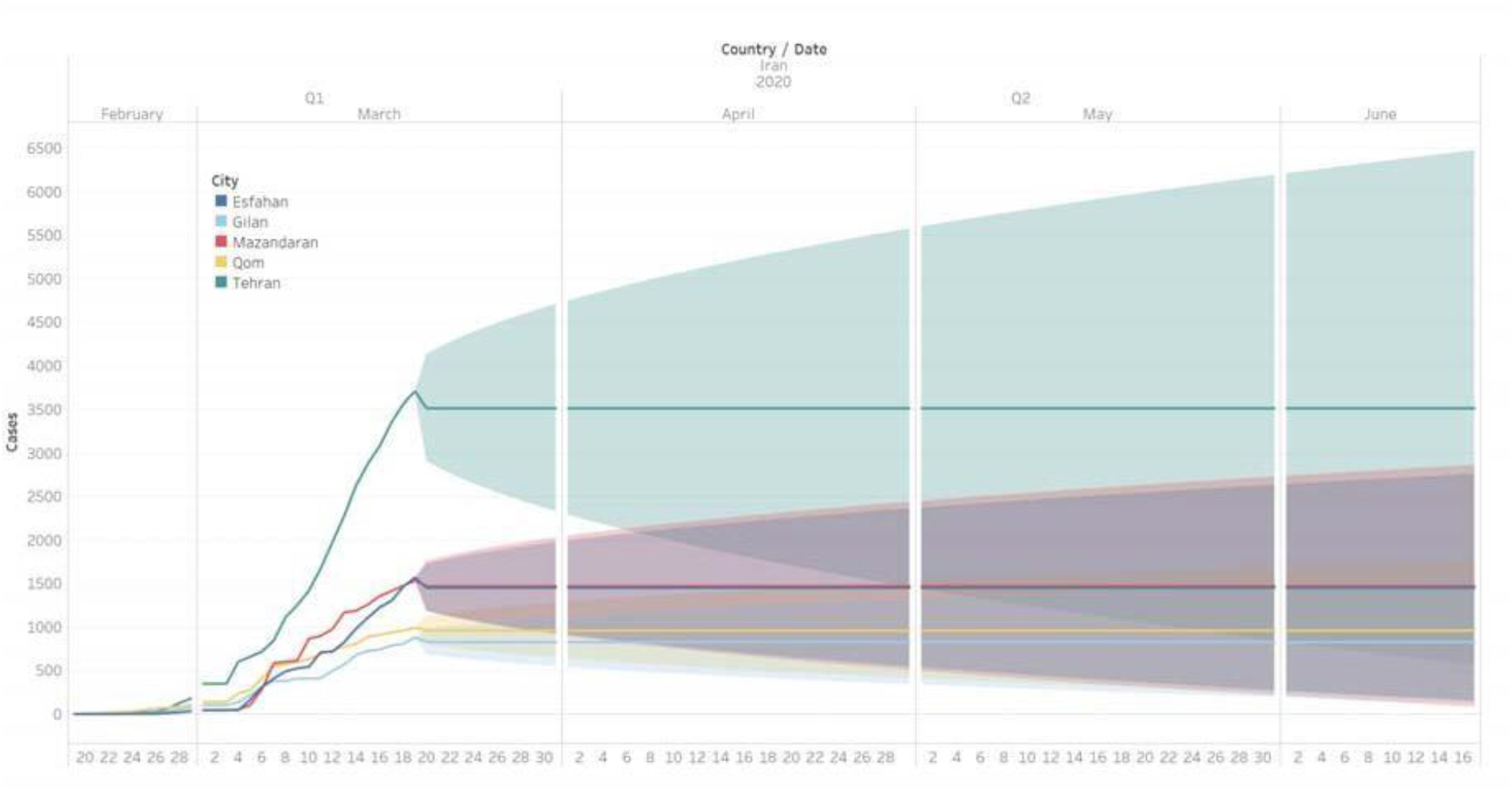

Two models have been tested with varying degree of accuracy. The base model shown in Illustration 5 is the simple forecast model, without trend or seasonality built-in. It has the highest AIC out of the three models across the 5 sampled provinces, although it has a low MAPE, when compared to the other three statistical metrics like RMSE, MAE, MASE it yields the highest margin of error in the group.

The quality of the predictive model for Tehran is acceptable because the dataset is more robust compared to one integrated with seasonality. The Mean Absolute Scaled Error, MASE is less than 1 for the projected 90 days period which is a positive score for the model. The RMSE, unbiased forecast score is slightly higher than the MAE, which gives a median future distribution of 84. The MAPE is over 100% which means the errors are “much greater” then the actual values. Table 1 & 2

**Table 1.**
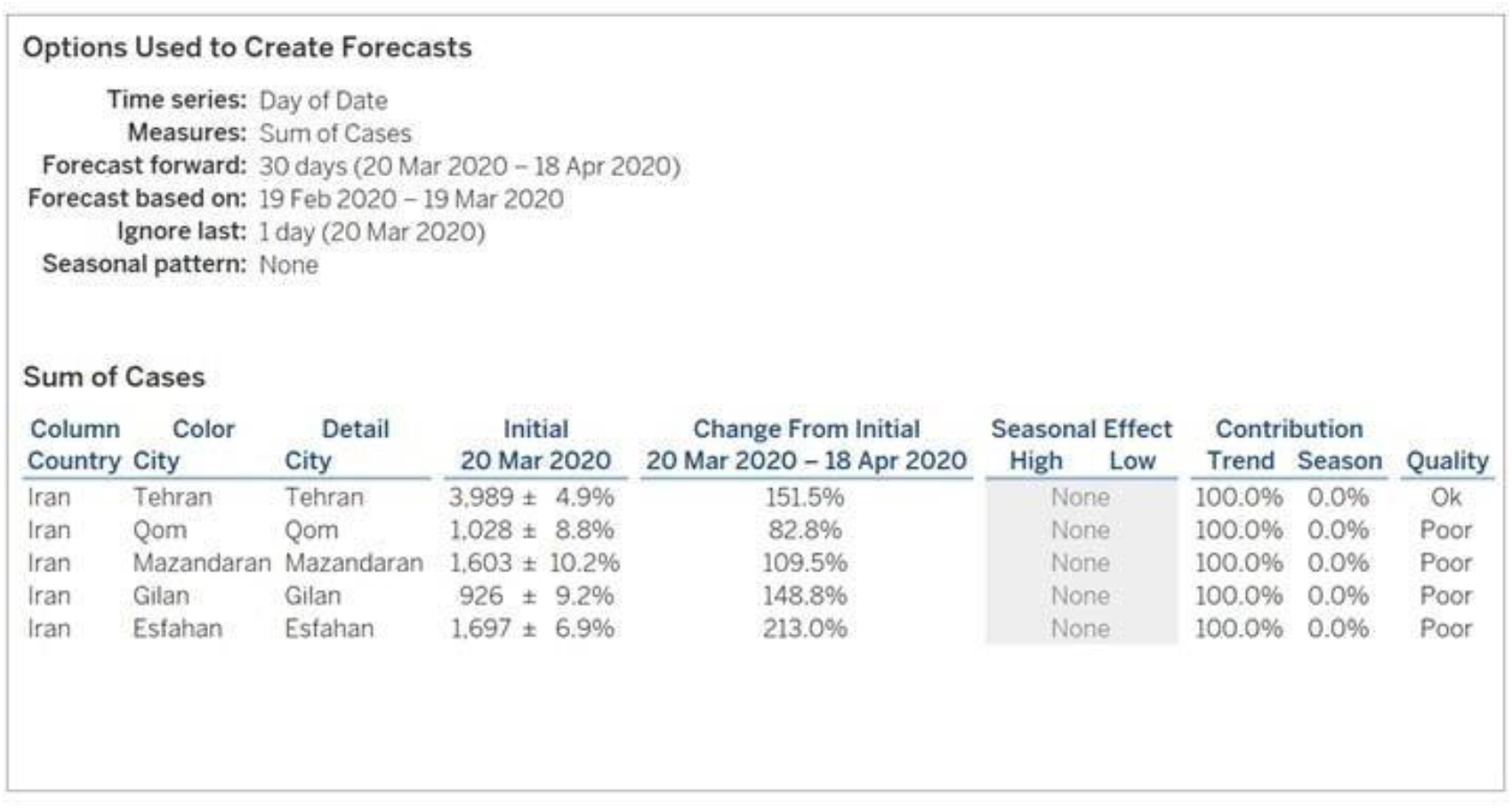

**Table 2.**
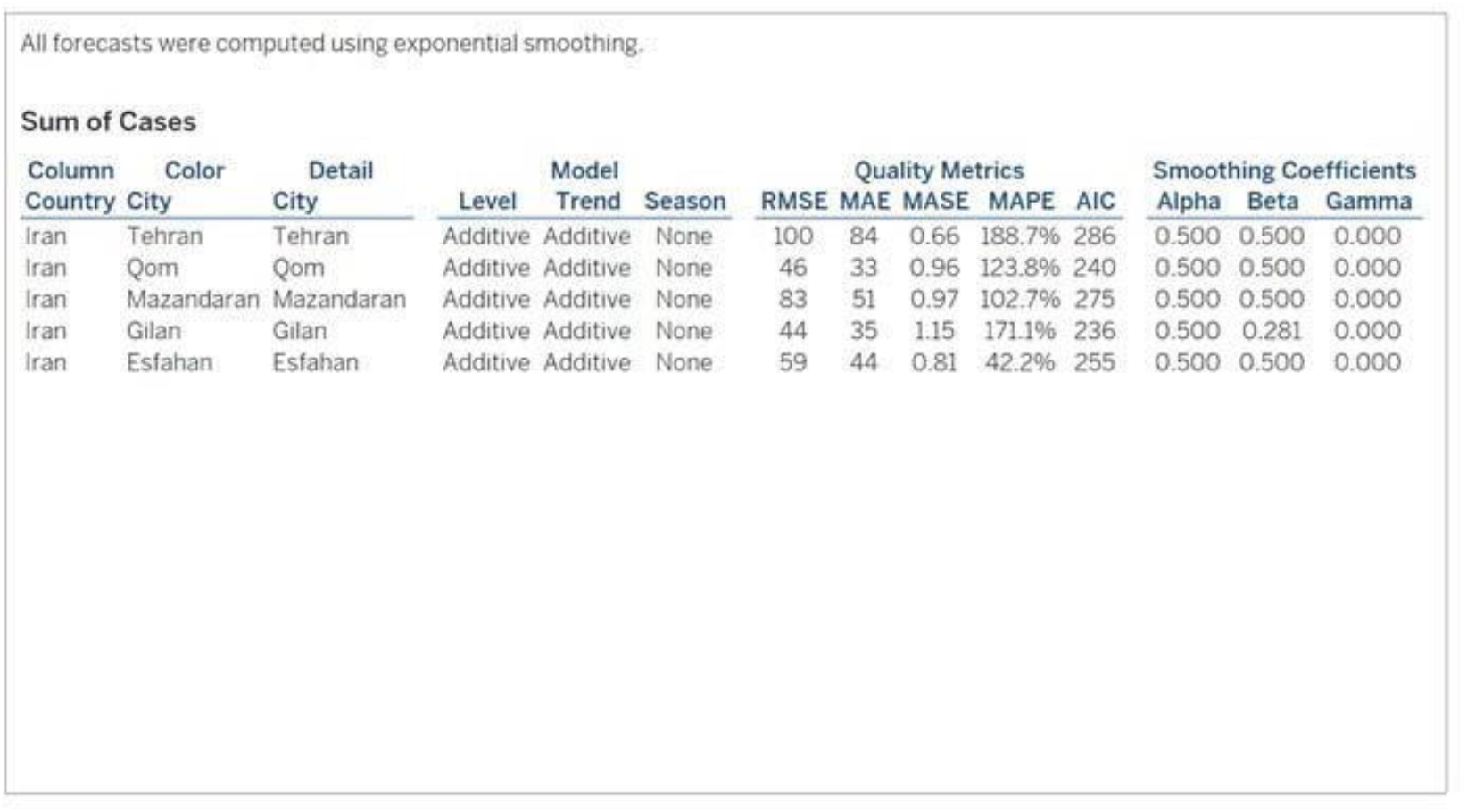

Sum of Cases
| Column | Color | Detail | Model |  |  | Quality Metrics |  |  |  |  | Smoothing Coefficients |  |  |
| --- | --- | --- | --- | --- | --- | --- | --- | --- | --- | --- | --- | --- | --- |
| Country | City | City | Level | Trend | Season | RMSE | MAE | MASE | MAPE | AIC | Alpha | Beta | Gamma |
| Iran | Tehran | Tehran | Additive | Additive | None | 100 | 84 | 0.66 | 188.7% | 286 | 0.500 | 0.500 | 0.000 |
| Iran | Qom | Qom | Additive | Additive | None | 46 | 33 | 0.96 | 123.8% | 240 | 0.500 | 0.500 | 0.000 |
| Iran | Mazandaran | Mazandaran | Additive | Additive | None | 83 | 51 | 0.97 | 102.7% | 275 | 0.500 | 0.500 | 0.000 |
| Iran | Gilan | Gilan | Additive | Additive | None | 44 | 35 | 1.15 | 171.1% | 236 | 0.500 | 0.281 | 0.000 |
| Iran | Esfahan | Esfahan | Additive | Additive | None | 59 | 44 | 0.81 | 42.2% | 255 | 0.500 | 0.500 | 0.000 |

**Iluustration 7.**
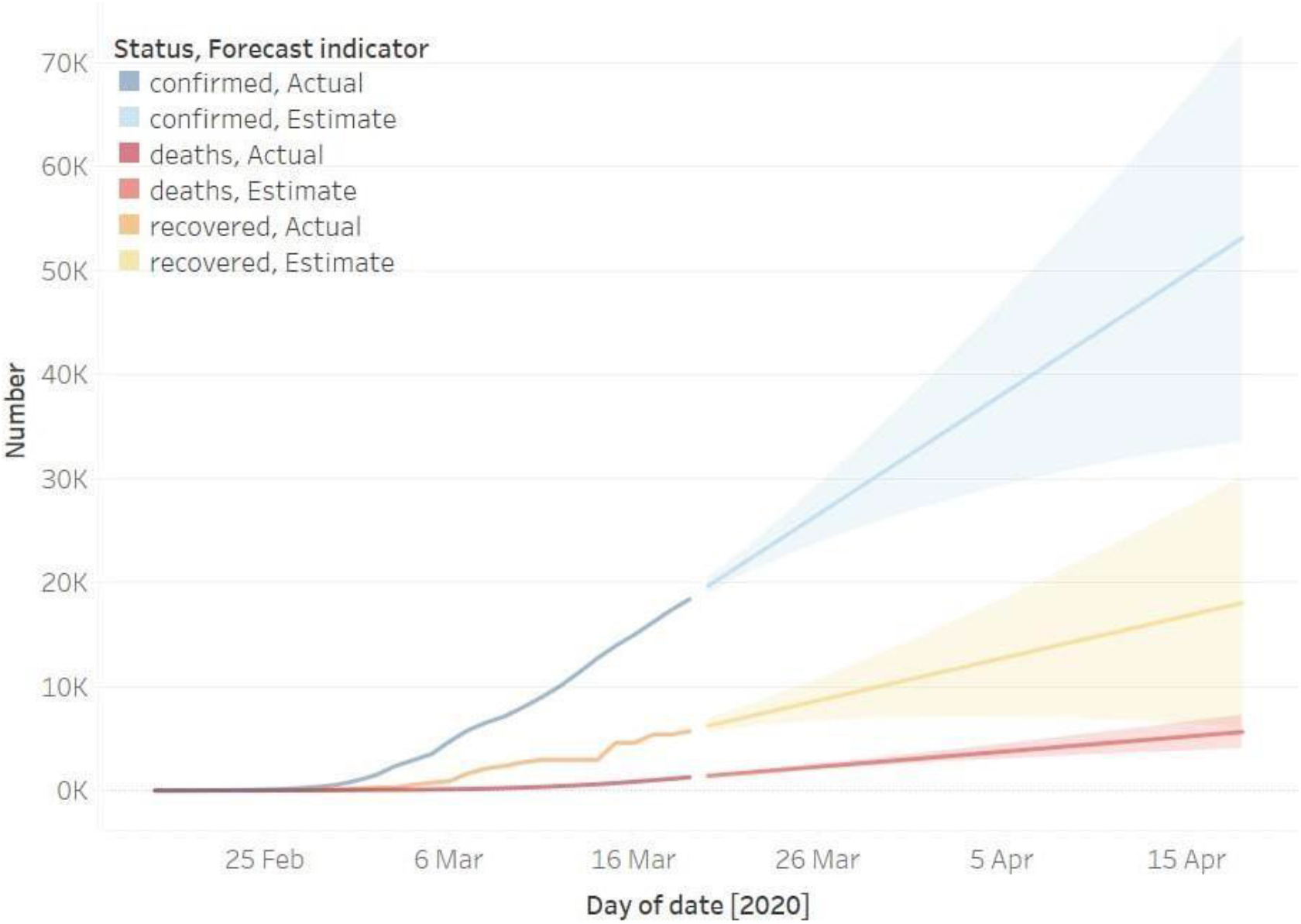

**Table 3.**
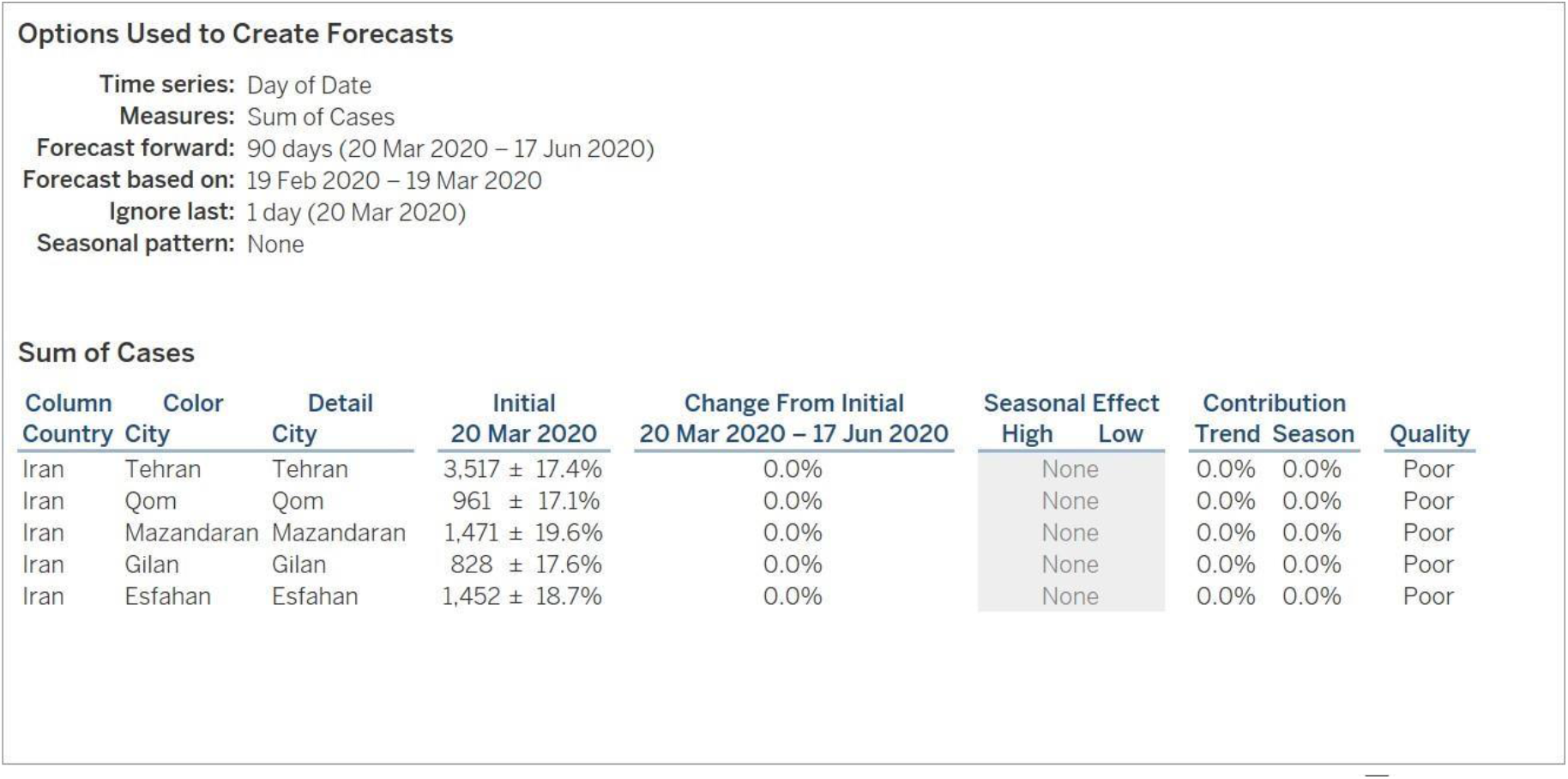

Sum of Cases
| Column | Color | Detail | Initial | Change From Initial | Seasonal Effect |  | Contribution |  |  |
| --- | --- | --- | --- | --- | --- | --- | --- | --- | --- |
| Country | City | City | 20 Mar 2020 | 20 Mar 2020 – 17 Jun 2020 | High | Low | Trend | Season | Quality |
| Iran | Tehran | Tehran | 3,517 ± 17.4% | 0.0% | None |  | 0.0% | 0.0% | Poor |
| Iran | Qom | Qom | 961 ± 17.1% | 0.0% | None |  | 0.0% | 0.0% | Poor |
| Iran | Mazandaran | Mazandaran | 1,471 ± 19.6% | 0.0% | None |  | 0.0% | 0.0% | Poor |
| Iran | Gilan | Gilan | 828 ± 17.6% | 0.0% | None |  | 0.0% | 0.0% | Poor |
| Iran | Esfahan | Esfahan | 1,452 ± 18.7% | 0.0% | None |  | 0.0% | 0.0% | Poor |

**Table 4.**

Sum of Cases
| Column<br>Country | Color<br>City | Detail<br>City | Model |  |  | Quality Metrics |  |  |  |  | Smoothing Coefficients |  |  |
| --- | --- | --- | --- | --- | --- | --- | --- | --- | --- | --- | --- | --- | --- |
|  |  |  | Level | Trend | Season | RMSE | MAE | MASE | MAPE | AIC | Alpha | Beta | Gamma |
| Iran | Tehran | Tehran | Additive | None | None | 313 | 235 | 1.84 | 33.8% | 351 | 0.500 | 0.000 | 0.000 |
| Iran | Qom | Qom | Additive | None | None | 84 | 64 | 1.88 | 37.7% | 272 | 0.500 | 0.000 | 0.000 |
| Iran | Mazandaran | Mazandaran | Additive | None | None | 147 | 98 | 1.85 | 33.2% | 306 | 0.500 | 0.000 | 0.000 |
| Iran | Gilan | Gilan | Additive | None | None | 75 | 55 | 1.82 | 30.7% | 265 | 0.500 | 0.000 | 0.000 |
| Iran | Esfahan | Esfahan | Additive | None | None | 138 | 97 | 1.79 | 30.8% | 302 | 0.500 | 0.000 | 0.000 |

The same process has been applied to review the confirmed, deaths and recovered dataset. The forecast has been carried out for the next 30 days, a shorter timeframe has been selected as there is a high probability that the Iranian New Year’s celebration, Farvardin, first month of Spring (30^th^ March in Western calendar) will have an impact on the infection rate following the event.

The best predictive model as seen in Illustration 7 predicts the confirmed cases to be in the range of 35,000-70,000, with the number of reported COVDI-19 deaths to be between 3,000 – 5,000 and 5,000 – 30,000 of recovered cases.

**Table 5.**

| Options Used to Create Forecasts |  |  |  |  |  |  |  |
| --- | --- | --- | --- | --- | --- | --- | --- |
| Time series: Day of date |  |  |  |  |  |  |  |
| Measures: Sum of Number |  |  |  |  |  |  |  |
| Forecast forward: 30 days (20 Mar 2020 – 18 Apr 2020) |  |  |  |  |  |  |  |
| Forecast based on: 19 Feb 2020 – 19 Mar 2020 |  |  |  |  |  |  |  |
| Ignore last: 1 day (20 Mar 2020) |  |  |  |  |  |  |  |
| Seasonal pattern: None (Searched for a seasonal pattern recurring every 7 Days) |  |  |  |  |  |  |  |
| Sum of Number |  |  |  |  |  |  |  |
| Color Status | Initial<br>20 Mar 2020 | Change From Initial<br>20 Mar 2020 – 18 Apr 2020 | Seasonal Effect<br>High Low |  | Contribution<br>Trend Season |  | Quality |
| recovered | 6,226 ± 11.5% | 189.9% | None |  | 100.0% | 0.0% | Poor |
| deaths | 1,411 ± 4.5% | 299.5% | None |  | 100.0% | 0.0% | Ok |
| confirmed | 19,661 ± 3.9% | 170.4% | None |  | 100.0% | 0.0% | Ok |

**Table 6.**

| All forecasts were computed using exponential smoothing. |  |  |  |  |  |  |  |  |  |  |  |
| --- | --- | --- | --- | --- | --- | --- | --- | --- | --- | --- | --- |
| Sum of Number |  |  |  |  |  |  |  |  |  |  |  |
| Color Status | Level | Model Trend | Season | Quality Metrics |  |  |  |  | Smoothing Coefficients |  |  |
|  |  |  |  | RMSE | MAE | MASE | MAPE | AIC | Alpha | Beta | Gamma |
| recovered | Additive | Additive | None | 364 | 257 | 1.31 | 15.9% | 364 | 0.500 | 0.314 | 0.000 |
| deaths | Additive | Additive | None | 32 | 26 | 0.59 | 240.7% | 218 | 0.500 | 0.500 | 0.000 |
| confirmed | Additive | Additive | None | 393 | 319 | 0.50 | 1,812.2% | 368 | 0.500 | 0.500 | 0.000 |

## Conclusion

Modeling outbreak ofCovid-19 shows that the number of patients and deaths is still increasing. Contagious diseases follow an exponential model and the same be Haves this one. This is because, the virus can spread to others and finally each person turns into a carrier of the virus and transmit it to another person. Disease control depends on disconnection and social distancing. In addition, many factors are effective in stopping the disease.

These include citizens’ participation in the prevention process, health education, the effectiveness of instructive traditions, environmental conditions, and so on. This article strived to analyze the growth trend of the number of patients, deaths and patients recovered in some provinces of Iran. The knowledge gained can help health planners and planners. Combining the findings of this study with other countries’ studies can help to extract a global pattern for the virus outbreak process analysis. We believe that there are still many factors that can be included in the study. Adding these factors helps to validate and consolidate the findings. We hope that our study as part of an effort to better understand the disease and prevent the spread of the disease will help a global achievement

## Data Availability

The data that support the findings of this study are available from the corresponding author, upon reasonable request.

## Notes

### Competing Interest Statement

The authors have declared no competing interest.

### Funding Statement

No funding body

### Summary of Updates

Due to the non-participation of Mrs. Mila MalekolKalami in the research process and her lack of cooperation, her name was removed from the list of authors in order to comply with the standards of research ethics.

